# Sex-Specific Incidence of Hemorrhagic Stroke in Japan: An Integrated Population-Based Study

**DOI:** 10.64898/2026.11.09.26367684

**Authors:** Satoshi Shitara, Akiko Harada, Sachiko Tanaka-Mizuno, Naoyuki Takashima, Takako Fujii, Hisatomi Arima, Naomi Miyamatsu, Makoto Urushitani, Kazuhiko Nozaki, Katsuyuki Miura, Kazumichi Yoshida

## Abstract

**Background:** Intracerebral hemorrhage (ICH) and subarachnoid hemorrhage (SAH) have contrasting epidemiological characteristics, including sex-specific incidence. However, their overall contribution to sex differences in hemorrhagic stroke remains unclear because they are generally evaluated separately. We examined the sex-specific epidemiology of hemorrhagic stroke by integrating ICH and SAH.

**Methods:** We analyzed first-ever ICH and SAH events recorded from 2011 through 2015 in the Shiga Stroke and Heart Attack Registry, a population-based registry covering approximately 1.4 million residents in Japan. Age-specific and age-adjusted incidence rates were calculated by sex and stroke subtype using direct standardization to the 2015 Japanese Census population.

**Results:** Among 4,517 patients with hemorrhagic stroke, 3,461 (76.6%) had ICH and 1,056 (23.4%) had SAH. The age-adjusted incidence of ICH was higher in men than in women (61.2 vs 49.8 per 100,000 person-years), whereas SAH incidence was more than twice as high in women as in men (22.4 vs 10.4 per 100,000 person-years). Despite these opposing patterns, the age-adjusted incidence of hemorrhagic stroke was nearly identical between men and women (71.6 vs 72.2 per 100,000 person-years). ICH accounted for a greater share of the overall incidence among men, whereas SAH accounted for a greater share among women.

**Conclusions:** Despite marked subtype-specific sex differences, the overall incidence of hemorrhagic stroke was similar between women and men. These findings demonstrate that substantial sex differences in individual subtypes may be offset when ICH and SAH are considered together, highlighting the value of an integrated population-level perspective in evaluating sex disparities in hemorrhagic stroke.

## Introduction

Intracerebral hemorrhage (ICH) and subarachnoid hemorrhage (SAH) are the major subtypes of hemorrhagic stroke and remain important causes of death and disability worldwide.^1,2^ Although both are classified as hemorrhagic stroke, their epidemiological characteristics differ substantially, including age at onset, risk-factor profiles, and sex distribution.^3^ These differences are clinically and biologically relevant because they may influence the overall population burden of hemorrhagic stroke in different ways.

Sex differences have been consistently reported for both ICH and SAH, but in opposite directions. ICH generally occurs more frequently in men than in women, whereas SAH is more common among women, particularly at older ages.^3–5^ These contrasting patterns have been investigated extensively within each stroke subtype.^6,7^ However, ICH and SAH have generally been evaluated as separate entities in epidemiological studies, and the implications of their opposing sex-specific distributions for the overall burden of hemorrhagic stroke remain unclear.

From a population perspective, evaluating these subtypes separately may provide an incomplete picture of sex disparities in hemorrhagic stroke. The higher incidence of ICH among men may be counterbalanced by the higher incidence of SAH among women, potentially resulting in a different overall sex-specific pattern of hemorrhagic stroke when the two subtypes are considered together. Thus, examining the major hemorrhagic stroke subtypes together may provide a complementary perspective on the population-level epidemiology of hemorrhagic stroke.

In this population-based study, we used data from the Shiga Stroke and Heart Attack Registry (SSHR) to characterize sex-specific differences in the incidence of ICH and SAH and to examine their combined contribution to the overall incidence of hemorrhagic stroke. We further examined whether the marked sex differences observed for ICH and SAH persisted when the two subtypes were considered together.

## Methods

### Study Design and Data Source

We conducted a population-based observational study using data from the SSHR. The SSHR is a population-based registry established to ascertain the incidence and clinical characteristics of acute stroke and acute myocardial infarction among residents of Shiga Prefecture, Japan.^6–8^ Shiga Prefecture has a population of approximately 1.4 million and comprises both urban and rural communities, providing a representative population for epidemiological assessment.

The SSHR systematically collects information on acute stroke events from participating medical institutions, including hospitals providing emergency and acute stroke care. Cases are identified through multiple sources, including hospital records and diagnostic information, to maximize case ascertainment. For the present study, we used SSHR data from 2011 through 2015.

### Study Population

We identified all patients with first-ever ICH or SAH who were residents of Shiga Prefecture and experienced an acute stroke event between January 1, 2011, and December 31, 2015.

ICH was defined as spontaneous bleeding into the brain parenchyma confirmed by neuroimaging or clinical diagnosis. SAH was defined as spontaneous subarachnoid bleeding confirmed by neuroimaging, cerebrospinal fluid examination, or clinical diagnosis. Traumatic hemorrhage was excluded.

For the present analysis, ICH and SAH were considered the major subtypes of hemorrhagic stroke and were analyzed both separately and in combination. Thus, the overall hemorrhagic stroke population comprised all eligible first-ever ICH and SAH events.

### Population at Risk and Incidence Rates

Population denominators were obtained from official population statistics for Shiga Prefecture during the study period. Person-years at risk were calculated according to sex and age group.

Age-specific incidence rates were calculated for ICH, SAH, and hemorrhagic stroke overall and were expressed as cases per 100,000 person-years. Age-adjusted incidence rates were calculated by the direct standardization method using the age-specific population distribution of the 2015 Japanese Census as the standard population.

For the integrated analysis, ICH and SAH events were combined to calculate the overall incidence of hemorrhagic stroke.

### Clinical Characteristics

Baseline demographic and clinical characteristics were obtained from the SSHR. For ICH, these included age, sex, Japan Coma Scale (JCS) score at presentation, hypertension, diabetes mellitus, history of coronary heart disease, antithrombotic therapy, smoking, alcohol consumption, and modified Rankin Scale (mRS) score at discharge.

For SAH, clinical characteristics included age, sex, World Federation of Neurosurgical Societies (WFNS) grade, hypertension, diabetes mellitus, history of coronary heart disease, smoking, alcohol consumption, and mRS score at discharge.

These characteristics were summarized according to sex for ICH and SAH separately.

### Statistical Analysis

Continuous variables are presented as mean ± standard deviation (SD) and categorical variables as numbers and percentages. Continuous variables were compared using one-way analysis of variance (ANOVA), and categorical variables were compared using the chi-square test or Fisher’s exact test, as appropriate.

Age-specific and age-adjusted incidence rates were calculated separately for ICH and SAH and for hemorrhagic stroke overall. The contribution of each subtype to the overall incidence of hemorrhagic stroke was examined across age groups and by sex.

All statistical analyses were performed using SAS version 9.4 (SAS Institute, Cary, NC). Two-sided P-values were reported for all analyses. A P-value <0.05 was considered significant.

## Results

### Study population

Between 2011 and 2015, a total of 4,517 patients with first-ever hemorrhagic stroke were identified in the SSHR, including 3,461 patients with ICH and 1,056 with SAH. Among patients with ICH, 1,894 (54.7%) were men and 1,567 (45.3%) were women, whereas among patients with SAH, 334 (31.6%) were men and 722 (68.4%) were women.

In both subtypes, women were older than men. Among patients with ICH, the mean age was 75±14 years in women and 69±14 years in men (*P*<0.001). Among patients with SAH, the corresponding values were 68±15 and 60±15 years, respectively (*P*<0.001).

### Baseline characteristics of ICH and SAH according to sex

Baseline characteristics of patients with ICH and SAH according to sex are shown in Tables 1 and 2, respectively. Among patients with ICH, women were older than men and had lower prevalences of diabetes mellitus, smoking, and alcohol consumption. The distributions of JCS and mRS scores also differed significantly between sexes. Among patients with SAH, women were older and had a higher prevalence of hypertension, whereas smoking and alcohol consumption were more common among men. The distributions of WFNS grade and mRS score did not differ significantly between sexes.

**Table 1.**
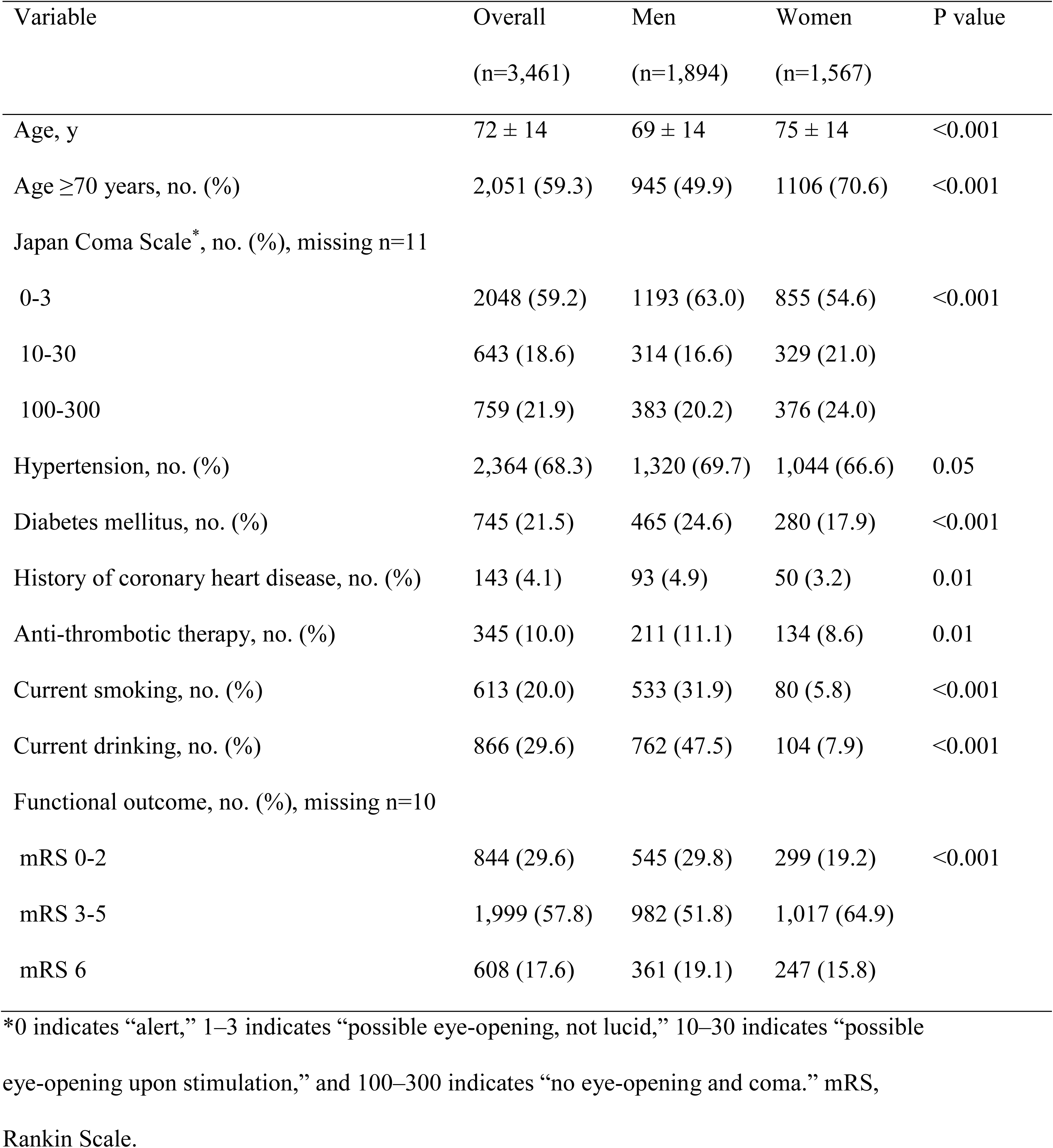
Baseline characteristics of intracerebral hemorrhage patients according to sex (N=3,461): Shiga Stroke and Heart Attack Registry, Shiga, Japan, 2011 to 2015.

**Table 2.** Baseline characteristics of subarachnoid hemorrhage patients according to sex (N=1,056): Shiga Stroke and Heart Attack Registry, Shiga, Japan, 2011 to 2015.

| Variable | Overall<br>(n=1,056) | Men<br>(n=334) | Women<br>(n=722) | P value |
|---|---|---|---|---|
| Age, y | 66 ± 15 | 60 ± 15 | 68 ± 15 | <0.001 |
| Age ≥70 years, no. (%) | 456 (43.2) | 94 (28.1) | 362 (50.1) | <0.001 |
| WFNS Grade, no. (%), missing n=8 |  |  |  |  |
| I-II | 454 (43.4) | 153 (46.2) | 301 (42.0) | 0.243 |
| III | 60 (5.7) | 20 (6.0) | 40 (5.6) |  |
| IV-V | 534 (50.9) | 158 (47.6) | 376 (52.4) |  |
| Hypertension, no. (%) | 588 (55.7) | 170 (50.9) | 418 (57.9) | 0.033 |
| Diabetes mellitus, no. (%) | 239 (22.6) | 85 (25.5) | 154 (21.3) | 0.137 |
| History of coronary heart disease, no. (%) | 39 (3.7) | 16 (4.8) | 23 (3.2) | 0.199 |
| Current smoking, no. (%) | 248 (27.1) | 142 (49.0) | 106 (16.9) | <0.001 |
| Current drinking, no. (%) | 228 (26.2) | 153 (54.3) | 75 (12.7) | <0.001 |
| Functional outcome, no. (%), missing n=5 |  |  |  |  |
| mRS 0-2 | 390 (46.9) | 138 (41.4) | 252 (34.9) | 0.101 |
| mRS 3-5 | 336 (31.8) | 101 (30.2) | 235 (32.5) |  |
| mRS 6 | 325 (30.8) | 92(27.5) | 233 (32.3) |  |

mRS indicates modified Rankin Scale; WFNS, World Federation of Neurosurgical Societies.

### Age-Specific Incidence of ICH and SAH

Age-specific incidence rates of ICH and SAH according to sex are shown in Figure 1. The incidence of ICH increased markedly with age in both sexes and was generally higher in men than in women, particularly at older ages. In contrast, SAH showed an opposingpattern, with higher incidence among women than men particularly from middle age onward.

**Figure 1.**
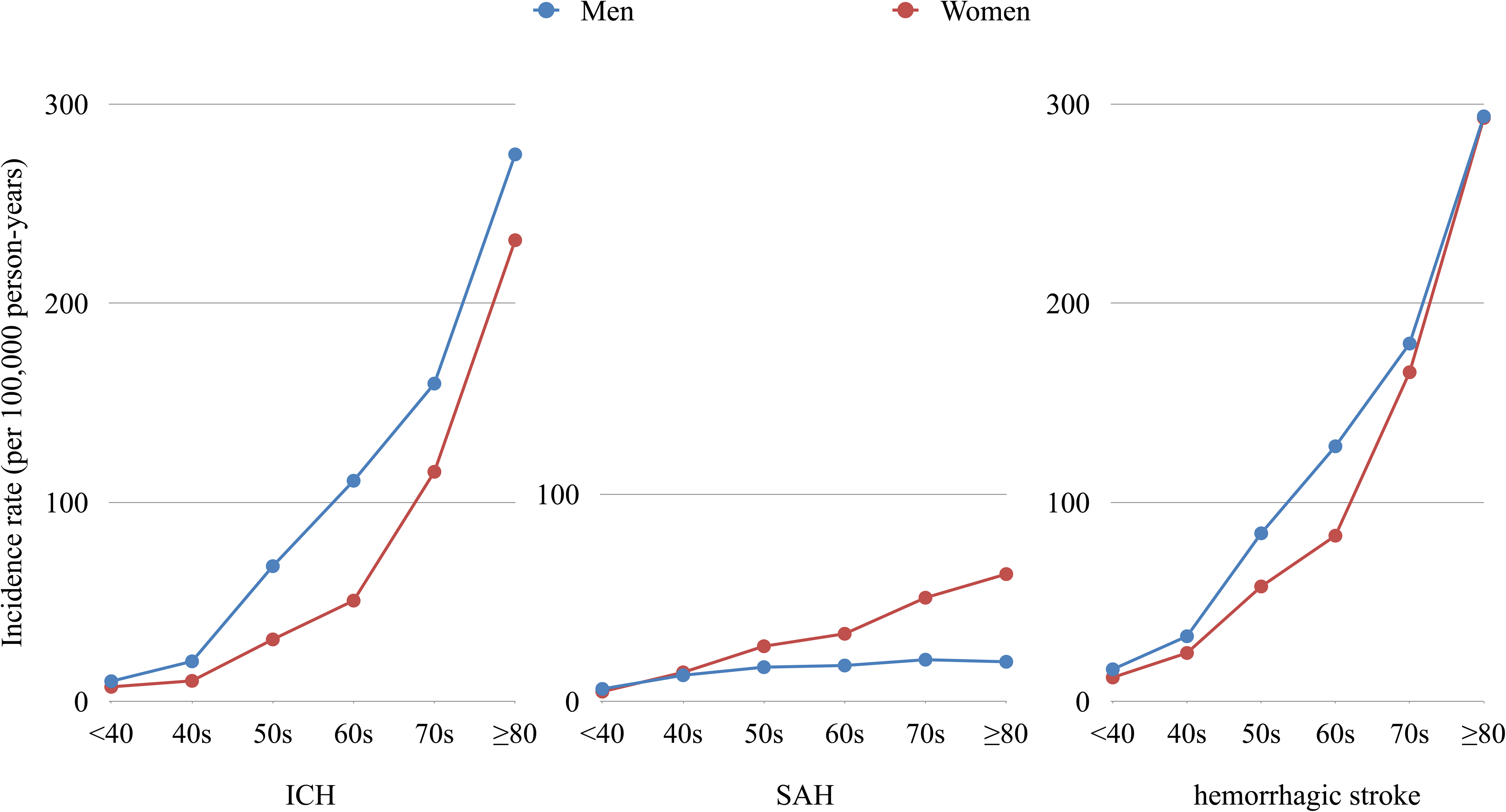
Age-specific incidence rates of hemorrhagic stroke subtypes according to sex. Age-specific incidence rates of intracerebral hemorrhage (ICH) (A), subarachnoid hemorrhage (SAH) (B), and overall hemorrhagic stroke (ICH and SAH combined) (C) among men and women. ICH incidence was higher in men, whereas SAH incidence was higher in women. When the two subtypes were combined, the age-specific incidence of overall hemorrhagic stroke was similar between men and women.

Using direct standardization based on the 2015 Japanese Census population, the age-adjusted incidence of ICH was 61.2 per 100,000 person-years in men and 49.8 per 100,000 person-years in women. The corresponding age-adjusted incidence of SAH was 10.4 per 100,000 person-years in men and 22.4 per 100,000 person-years in women.

### Integrated Burden of Hemorrhagic Stroke

When ICH and SAH were considered together, the age-adjusted incidence of hemorrhagic stroke was 71.6 per 100,000 person-years in men and 72.2 per 100,000 person-years in women, indicating a nearly identical overall incidence between sexes (Figure 2). The higher incidence of ICH among men was largely offset by the higher incidence of SAH among women. Specifically, the male excess in ICH incidence was 11.4 per 100,000 person-years, whereas the female excess in SAH incidence was 12.0 per 100,000 person-years.

**Figure 2.**
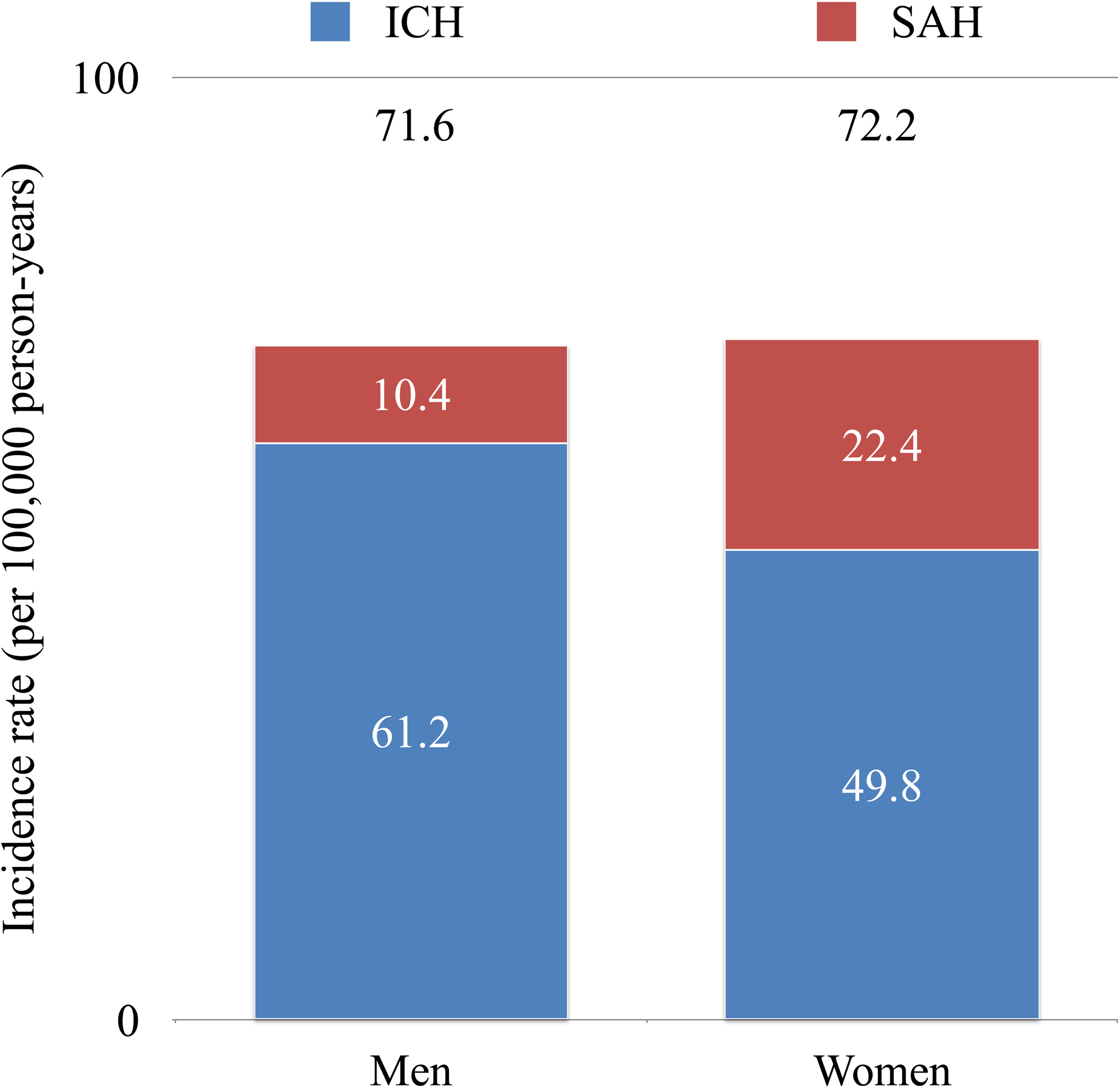
Contribution of intracerebral hemorrhage and subarachnoid hemorrhage to overall hemorrhagic stroke incidence according to sex. Stacked bars show the age-adjusted incidence rates of intracerebral hemorrhage (ICH) and subarachnoid hemorrhage (SAH), expressed as cases per 100,000 person-years. Men had a higher incidence of ICH than women, whereas women had a substantially higher incidence of SAH. These opposing subtype-specific patterns resulted in a similar overall incidence of hemorrhagic stroke between men and women (71.6 vs 72.2 per 100,000 person-years, respectively).

These opposing contributions resulted in a similar overall incidence of hemorrhagic stroke despite substantial differences in subtype-specific incidence between sexes. As shown in Figure 1, ICH incidence was higher among men, particularly at older ages, whereas SAH incidence was higher among women from middle age onward.

## Discussion

### Principal Findings

In this population-based study, we found that the incidence of ICH was higher in men than in women, whereas the incidence of SAH was more than twice as high in women as in men. Despite these marked and opposing subtype-specific differences, the age-adjusted incidence of hemorrhagic stroke was nearly identical between men and women. These findings demonstrate that substantial sex differences at the subtype level can be offset when the major hemorrhagic stroke subtypes are considered together, resulting in a similar overall incidence between sexes.

### Opposing Sex-Specific Patterns of ICH and SAH

The higher incidence of ICH among men observed in our study is consistent with previous population-based studies.^6,9,10^ Several factors may contribute to this sex difference, including differences in the prevalence and control of vascular risk factors and lifestyle-related exposures. In our cohort, men had higher prevalences of smoking, alcohol consumption, and diabetes mellitus than women. Differences in vascular risk factors and lifestyle-related exposures may partly contribute to the sex difference in ICH incidence.^10^ The higher age-adjusted incidence of ICH in men, despite the older age of womenin our cohort, further highlights the sex-specific epidemiological pattern of ICH.

In contrast, SAH showed the opposite pattern, with a substantially higher incidence among women. The female predominance of SAH has been consistently reported in population-based studies and may reflect a combination of age-related and sex-specific biological factors.^7,8,11^ In our cohort, women with SAH were considerably older than men, and the female predominance became more apparent from middle age onward. Hormonal and vascular changes associated with aging and menopause have been proposed as potential contributors to the higher incidence of SAH among women, although the mechanisms underlying this sex difference remain incompletely understood.^12^

### A Different Perspective on the Population Burden of Hemorrhagic Stroke

The principal finding of the present study was that the opposing sex-specific patterns of ICH and SAH resulted in an almost identical overall incidence of hemorrhagic stroke between men and women. The age-adjusted incidence of ICH was 11.4 per 100,000 person-years higher in men than in women, whereas the incidence of SAH was 12.0 per 100,000 person-years higher in women than in men. Thus, the excess incidence of ICH among men was largely offset by the excess incidence of SAH among women.

This finding provides a different perspective on sex disparities in hemorrhagic stroke. Previous epidemiological studies have described sex differences in ICH and SAH, reflecting their distinct epidemiological and clinical characteristics.^3^ However, evaluating these subtypes separately may provide an incomplete picture of the overall population burden of hemorrhagic stroke. Our findings demonstrate that marked sex differences in individual stroke subtypes do not necessarily translate into a corresponding sex difference in the overall incidence of hemorrhagic stroke.

Importantly, this observation should not be interpreted as indicating that men and women have similar epidemiological risk profiles. Rather, the similar overall incidence reflects differences in the subtype composition of hemorrhagic stroke between the sexes. Although ICH was the predominant subtype in both sexes, it accounted for a greater share of the overall incidence among men, whereas SAH accounted for a greater share among women. Thus, the apparent similarity in overall incidence conceals substantial differences in the composition of hemorrhagic stroke between men and women.

### Implications for Stroke Epidemiology and Prevention

These findings have implications for how the population burden of hemorrhagic stroke is evaluated. When disease burden is assessed using a broad stroke category, substantial differences between component subtypes may be masked. Conversely, analyses focusing exclusively on individual subtypes may not fully capture the overall pattern of sex differences in hemorrhagic stroke.

An integrated approach that considers the major hemorrhagic stroke subtypes together may therefore provide complementary information to conventional subtype-specific analyses. This perspective may be particularly relevant to population-level prevention planning, because the relative contributions of ICH and SAH to the overall burden of hemorrhagic stroke differ between men and women. In men, the greater contribution of ICH highlights the importance of addressing established risk factors for ICH, whereas in women, the greater contribution of SAH underscores the importance of SAH-specific prevention efforts.^4,13^

### Strengths and Limitations

Our study has several strengths. First, the use of a population-based stroke registry enabled assessment of the incidence of both ICH and SAH within the same underlying population using a consistent case-ascertainment framework. Second, the relatively large number of hemorrhagic stroke events allowed direct comparison of sex-specific incidence patterns across the two major subtypes. Third, applying the same age-standardization approach to both subtypes and their combined incidence enabled direct assessment of their relative contributions to the overall burden of hemorrhagic stroke.

Several limitations should be considered. First, the study period was limited to 2011 through 2015, and the epidemiology of hemorrhagic stroke may have changed since then because of changes in population structure, vascular risk-factor management, and treatment patterns. Second, although the registry covered a large population, the findings were derived from a single prefecture in Japan and may not be directly generalizable to other populations with different demographic or risk-factor profiles. Third, the present study was designed primarily to describe population-level incidence and did not investigate the mechanisms underlying the opposing sex-specific patterns of ICH and SAH. Finally, because ICH and SAH have distinct pathophysiological mechanisms, their integration in the present study should be interpreted as an assessment of the overall population burden of hemorrhagic stroke rather than as an indication that they constitute a biologically homogeneous disease entity.

## Conclusion

In this population-based study, ICH and SAH demonstrated opposing sex-specific incidence patterns, with ICH more common in men and SAH more common in women. However, when these two major subtypes were considered together, the overall incidence of hemorrhagic stroke was nearly identical between sexes. These findings indicate that the overall sex-specific incidence of hemorrhagic stroke reflects the contrasting contributions of its major subtypes and highlight the value of an integrated population-level perspective in evaluating sex disparities in hemorrhagic stroke.

## Data Availability

The data that support the findings of this study are not available on the ethical grounds. Further inquiries can be directed to the corresponding author.

## Acknowledgments

The authors would like to thank all investigators, participating hospitals, and study staff at the Shiga Stroke Data Center, Shiga Medical Association, Shiga Prefecture Hospital Association, Biwako Brain & Heart Attack Consortium, Department of Public Health Care and Welfare, and the Shiga Prefectural government.

## Sources of Funding

The Shiga Stroke and Heart Attack Registry is supported by Shiga Prefecture and the Japan Agency for Medical Research and Development (Grant No. 17ek0210090). The funding sources listed above had no role in the study design, collection, analyses, and interpretation of the results.

## Authors’ Contributions

SS designed the study protocol, analyzed data, and prepared the manuscript. AH, ST-M, NT, TF, HA, MU, KN, KM and KY advised the study planning, developed the theoretical content, interpreted the results, and approved the final version of the manuscript.

## Disclosures

None.

## Non-standard Abbreviations and Acronyms

ICH: intracerebral hemorrhage
mRS: modified Rankin Scale
SAH: subarachnoid hemorrhage
SSHR: Shiga Stroke and Heart Attack Registry
WFNS: World Federation of Neurosurgical Societies

## Notes

### Competing Interest Statement

The authors have declared no competing interest.

### Author Declarations

The study was approved by the Ethics Committee of Shiga University of Medical Science (approval number: 23-186-1).

## References

1. Xu L, Wang Z, Wu W, Li M, Li Q. Global, regional, and national burden of intracerebral hemorrhage and its attributable risk factors from 1990 to 2021: results from the 2021 Global Burden of Disease Study. BMC Public Health. 2024;24:2426.

2. GBD 2021 Global Subarachnoid Hemorrhage Risk Factors Collaborators; Rautalin I, Volovici V, Stark BA, Johnson CO, Kaprio J, Korja M, Krishnamurthi RV, Nair BS, Ranta A, et al. Global, Regional, and National Burden of Nontraumatic Subarachnoid Hemorrhage: The Global Burden of Disease Study 2021. JAMA Neurol. 2025;82:765–787.

3. Appelros P, Stegmayr B, Terént A. Sex differences in stroke epidemiology: a systematic review. Stroke. 2009;40:1082–1090.

4. Wang S, Zou XL, Wu LX, Zhou HF, Xiao L, Yao T, Zhang Y, Ma J, Zeng Y, Zhang L. Epidemiology of intracerebral hemorrhage: A systematic review and meta-analysis. Front Neurol. 2022;13:915813.

5. de Rooij NK, Linn FH, van der Plas JA, Algra A, Rinkel GJ. Incidence of subarachnoid haemorrhage: a systematic review with emphasis on region, age, gender and time trends. J Neurol Neurosurg Psychiatry. 2007;78:1365–1372.

6. Shitara S, Harada A, Tanaka-Mizuno S, Takashima N, Fujii T, Arima H, Urushitani M, Nozaki K, Miura K, Yoshida K. Contemporary Epidemiology of Intracerebral Hemorrhage in Japan: A Population-Based Study from the Shiga Stroke and Heart Attack Registry. Cerebrovasc Dis. Published online October 1, 2026.

7. Shitara S, Tanaka-Mizuno S, Takashima N, Fujii T, Arima H, Kita Y, Tsuji A, Kitamura A, Urushitani M, Miura K, et al. Population-Based Incidence Rates of Subarachnoid Hemorrhage in Japan: The Shiga Stroke and Heart Attack Registry. J Stroke. 2022;24:292–295.

8. Takashima N, Arima H, Kita Y, Fujii T, Miyamatsu N, Komori M, Sugimoto Y, Nagata S, Miura K, Nozaki K. Incidence, Management and Short-Term Outcome of Stroke in a General Population of 1.4 Million Japanese - Shiga Stroke Registry. Circ J 2017;81:1636–1646.

9. Foschi M, D’Anna L, Gabriele C, Conversi F, Gabriele F, De Santis F, Orlandi B, De Santis F, Ornello R, Sacco S. Sex Differences in the Epidemiology of Intracerebral Hemorrhage Over 10 Years in a Population-Based Stroke Registry. J Am Heart Assoc. 2024;13:e032595.

10. Wang S, Zou XL, Wu LX, Zhou HF, Xiao L, Yao T, Zhang Y, Ma J, Zeng Y, Zhang L. Epidemiology of intracerebral hemorrhage: A systematic review and meta-analysis. Front Neurol. 2022;13:915813.

11. Etminan N, Chang HS, Hackenberg K, de Rooij NK, Vergouwen MDI, Rinkel GJE, Algra A. Worldwide Incidence of Aneurysmal Subarachnoid Hemorrhage According to Region, Time Period, Blood Pressure, and Smoking Prevalence in the Population: A Systematic Review and Meta-analysis. JAMA Neurol. 2019;76:588–597.

12. Lai PMR, Jimenez M, Du R, Rexrode K. Association of Reproductive Life Span and Age at Menopause With the Risk of Aneurysmal Subarachnoid Hemorrhage. Neurology. 2022;98:e2005–e2012.

13. Feigin VL, Rinkel GJ, Lawes CM, Algra A, Bennett DA, van Gijn J, Anderson CS. Risk factors for subarachnoid hemorrhage: an updated systematic review of epidemiological studies. Stroke. 2005;36:2773–2780.

